# Understanding the Challenges of Medicine Optimisation Among Older People from Ethnic Minority Communities (Aged 60 Years and Above) With Polypharmacy in Primary Care: A Realist Review Protocol

**DOI:** 10.1101/2024.10.01.24314538

**Authors:** Nesrein Hamed, Clare Bates, Muhammed Umair Khan, Ian Maidment

**Affiliations:** Aston University; National Institute for Health and Care Research; University of Birmingham

**Keywords:** Multimorbidity, Person-centered care, Medication review, Medication management, Cultural competence, Deprescribing, Overprescribing, Under prescribing

## Abstract

**Background:** The number of older adults from ethnic minority communities (EMCs) in England and Wales particularly those aged 60 and above is increasing. This demographic change, which is usually coupled with the prevalence of polypharmacy among these populations presents unique challenges in the context of medicine optimisation. Failure in this context can lead to exacerbated health disparities, non-adherence, and inappropriate prescribing (whether over or under).

This review builds on the MEMORABLE study which was also a realist study that explored medication management in older people. This study aims to understand the complexities of medicine optimisation and what works and does not work, when and under what circumstances for older adults from EMCs. Key possible areas include cultural backgrounds, traditional beliefs, and systemic barriers that may influence health-seeking behaviours and medicine optimisation.

**Methods:** The review follows the five-step approach. Firstly, we will establish initial program theories to highlight the expected context, mechanisms, and outcomes. Following this, a formal search for evidence will be conducted. The third step involves the selection and appraisal of studies, studies will be screened by title, abstract/keywords and full text against inclusion and exclusion criteria. In the fourth stage, data from these studies will be extracted, recorded, and coded. The final step will synthesise this information, to test, refine, and expand our initial programme theories to understand how medicine optimisation works or does not work in these populations.

**Discussion:** This review will be conducted in line with the RAMESES reporting standards. This will include publishing the review in a scientific journal and submitting abstracts for presentation at both national and international primary care and pharmacy practice conferences. Once we improve the understanding of how medicine optimisation works for these populations with polypharmacy in primary care effective interventions can be developed.

**Systematic review registration:** PROSPERO registration number CRD42023432204

## Background

In recent years, there has been a significant increase in the number of older adults aged 60 years and above from ethnic minority communities (EMCs) living in England and Wales (1). The 2021 Census report found that the population aged 65 years and over was more ethnically diverse in 2021 than in 2011, with an increase from 4.5% to 6.4% in the older adults from EMCs aged 65 years and above (2). This rapid demographic ageing presents crucial challenges for these communities in the UK.

Previously, EMCs were collectively referred to as BAME, including various groups such as Black, Asian, and other minority ethnic origins, including Arab, Jewish, Gipsy, Roma, and Traveller communities (3). However, according to the ‘Commission on Race and Ethnic Disparities’ report published in March 2021, the term has faced criticism for its lack of specificity, its tendency to homogenise diverse experiences, and its inability to capture the diversities within ethnic groups (4). As a result, many institutions, organisations, and individuals in the UK have moved away from using BAME in favour of more specific and nuanced terms, such as specifying particular ethnic groups by name or using terms like ethnic minorities. (3, 4). In this review, the focus will be on a diverse range of ethnicities, including individuals of African, Caribbean, South Asian (Indian, Pakistani, Bangladeshi, Sri Lankan), Middle Eastern origins and individuals of mixed race. The religious diversity of these communities will also be considered. These populations were chosen to capture a wide range of experiences, which are influenced by diverse cultural, genetic, and socio-economic factors. Notably, they are among the largest ethnic communities in the UK. According to the 2021 Census, the white ethnic group remains the majority at 81.7% of the population in England and Wales. However, the Asian ethnic groups form the second largest category, making up 9.3% of the population, followed by Black ethnic groups at 4.0%, mixed ethnic groups at 2.9%, and other ethnic groups at 2.1% (5).

With age and associated multimorbidity, the use of five or more medication items by individuals (so-called polypharmacy) is more frequent (6). A study estimated that about 40% of people over 65 years take between five and nine medicines daily, while nearly 18% take 10 or more (7). The increase in medication items might lead to iatrogenic harm, with the likelihood escalating with both the patient‘s age and the quantity of prescribed medications (8). These include an increased risk of death, a higher likelihood of experiencing falls, adverse reactions to the drugs, long hospital stays, and a greater chance of being readmitted to the hospital shortly after being discharged (9, 10). In the UK, issues related to polypharmacy cause 5%–8% of unexpected hospital visits, costing the National Health Service (NHS) £530 million and leading to 5,700 deaths each year (11). These detrimental effects of polypharmacy are particularly challenging among older adults due to the physiological changes associated with ageing, such as impairment of metabolism and drug excretion, which could further induce drug-drug or drug-disease interactions (12). Moreover, cognitive impairment in this demographic exacerbates these challenges, complicating medicine optimisation and increasing the risk of medication errors (13).

Disproportionally, polypharmacy impacts older adults from EMCs (14). While the prevalence of polypharmacy is often associated with an aging population with multimorbidity, this explanation does not fully address the specific challenges faced by older adults from EMCs. Individuals from EMCs are influenced by cultural backgrounds that affect health-seeking behaviours and relationships with medications (15) and often have distinct health needs and expectations from healthcare systems compared to native populations (16). Their countries of origin may influence the supplementary use of healthcare services from abroad (16). Additionally, language barriers and the lack of support networks can worsen chronic conditions, which may lead to increased polypharmacy (17). Socioeconomic status further complicates this issue, with higher rates of polypharmacy in more deprived areas where EMCs are more likely to reside (14, 18).

The National Institute for Health and Care Excellence (NICE) define medicines optimisation (MO) as a “person-centred approach to safe and effective medicines use, to ensure people obtain the best possible outcomes from their medicines.“ (19). MO includes a range of strategies and interventions, including conducting medication reviews, deprescribing when necessary, performing medicine reconciliations, identifying potentially inappropriate prescriptions, and providing social support (20, 21). The failure to effectively optimise medicines within these contexts can lead to exacerbated health disparities, non-adherence, and inappropriate prescribing (whether over or under) (22).

MO especially within EMCs, faces a complex set of challenges that may lead to it not working effectively (23). Older adults from EMCs often bring a richness of traditional beliefs, past healthcare experiences, and potentially even systemic biases they have encountered (24). Such contexts can deeply influence their interactions within the healthcare system (25, 26). Through an evaluative lens, medicine optimisation is not just about prescribing and adherence; it‘s also a reflection of the balance between clinical needs and cultural layers. For instance, while a particular medication regimen may be clinically optimal, it may clash with traditional beliefs or past experiences, such as the reliance on herbal remedies or spiritual healing practices prevalent in some cultures (26). This clash can lead to non-adherence, as individuals may prioritise these long-standing cultural practices over prescribed medical treatments (26).

For patients from these communities, a primary barrier is often rooted in communication, where language disparities and cultural differences in transmitting health-related information, may prevent effective communication with healthcare practitioners (27). Health literacy goes beyond understanding prescription instructions, recognising potential side effects, and comprehending the implications of non-adherence to medication regimens (28-30). Given the cultural richness of EMCs, health directions and advice might need interpretation not just linguistically but also culturally (31, 32). What might seem like a straightforward directive in one culture could have different implications in another (32). This becomes even more vital when we consider traditional beliefs about health and illness that might differ significantly from contemporary medical perspectives (27).

Cultural competence is critical for healthcare practitioners, enabling them to effectively integrate patients‘ cultural beliefs and practices into medical care, thus fostering trust and adherence to treatments (31). However, the absence of cultural competence can lead to biases or stereotypes, which could negatively impact the outcome of the MO’s strategies and interventions (31). This is particularly important during the interactions between patient and practitioner in the decision-making process (33-35). Furthermore, practitioners face the challenge of integrating evidence-based medicine with traditional health beliefs. This complex task demands empathy and a deep understanding, which can be hard because of a lack of cultural competence training and support (36, 37). Additionally, systemic challenges, such as time constraints during consultations, workload, and pressure might impact the ability to engage deeply with patients’ unique contexts (38-40).

On a system level, healthcare structures may increase the disparities among these populations through policies that lack inclusiveness and cultural sensitivity (41). Unbalance between health policy directives and the unique needs of these populations may exacerbate barriers to effective MO (42). For instance, a system that prioritises digital communication and virtual consultations might disproportionately impact the older adults from EMCs who might be less digitally literate or have limited access to technological resources (42, 43).

The existing research on medicine optimisation for older adults in EMCs is notably limited, which highlights a significant gap in our understanding of how MO work for this population. While some studies have touched on specific challenges like communication barriers and social influence, few have holistically addressed the context of individual, familial, cultural, and systemic factors (44). The MEMORABLE study identified some of the medication-related challenges faced by these communities through a realist synthesis approach and critically the need for more empirical work (23, 45).

This realist review builds on the MEMORABLE study and work by others (45). With aims to develop a better understanding of how medicine optimisation works/does not work for older adults from EMCs, when and under what circumstances. This realist review will be guided by the following sub-questions:

- How does the context including cultural, socioeconomic, and healthcare system-related factors influence medicine optimisation?
- How do these contextual elements interact and drive the mechanisms to impact patient outcomes?
- What are the mechanisms that drive successful or unsuccessful medicine optimisation among older adults in EMCs?

## Method

This protocol adheres to Systematic Reviews and Meta-Analyses Protocols (PRISMA-P) guidelines (*Additional file 1*) (46). It is also registered in the International Prospective Register of Systematic Reviews (PROSPERO) under the registration number CRD42023432204.

### Overview

A Realist review aims to understand why interventions work or not, rather than summarising findings, by identifying underlying mechanisms and context (47-49). This type of review adopts a systematic, iterative process, grounded on the principle of understanding ‘what works, for whom, in what circumstances, and why’. It strategically explores the complexity of interventions and their varied outcomes across diverse contexts (47, 48).

This approach is centred around context, mechanism, and outcome configurations (CMOCs) (47). The ‘context‘ relates to the setting against which the medicine optimisation interventions occur (47). Context includes everything from societal norms and family structures to the nuances of the healthcare policies and primary care practices that shape their experiences. ‘Mechanism‘ refers to what prompts a change in the behaviour or circumstances of these patients (47). It could be the way information is communicated, and the trust between patient-practitioner. This interaction between context and mechanism is what makes healthcare services or interventions effective or ineffective in varying contexts (50). ‘Outcomes‘ extend beyond clinical results to include broader impacts such as improved understanding of medication, adherence to treatments, patient satisfaction, and overall quality of life (47).

Following a structured yet adaptable five-step framework as proposed by Pawson et al., the review process will include developing initial programme theories (IPTs), searching for evidence, selecting, and appraising relevant studies, and synthesising the data (51). This approach is not linear; steps may overlap or proceed concurrently, which could lead to adapting to new insights. In terms of rigorous standards of quality and reporting, this review will adhere to RAMESES guidelines (52). This adherence to the guideline will enhance the robustness and credibility of the conclusions, thereby contributing significantly to our understanding of how medicine optimisation works/does not for these populations (52).

### Step 1: Develop the Initial Programme Theories

Program theories play a vital role in guiding realist reviews (53). They are abstract representations that describe the components of interventions and lay down assumptions about how they lead to intended or observed outcomes. These theories explain the complex interaction between contexts, mechanisms, and outcomes, often represented as CMOCs (53).

We will develop ideas about IPTs from several sources, including empirical evidence, practitioners’ inputs, and formal theoretical frameworks. In this review, the Social Cognitive Theory (SCT) will be selected, which was developed by Bandura in 1986 as a theoretical framework (54). This theory is crucial in understanding the connection between individual behaviours, personal factors, and the social environment, which is particularly relevant in the context of medicine optimisation among older adults from EMCs (54). While other theories like the Health Belief Model and Theory of Planned Behaviour provide insights into health decisions (55, 56). However, they lack the focus on social and observational learning (57). The SCT looks into the interaction between personal factors, behaviours, and environment, which offers a deeper understanding of medicine optimisation among older adults from EMCs (58). Then, we will engage with practitioners and public and patient involvement (PPI) representatives to review and prioritise a maximum of 4 initial programme theories to test against the literature.

### Step 2: Develop the Search Strategy

Secondary data from academic and grey literature will be used to refine further the initial programme theory/theories. We will conduct an iterative literature search, with different search terms and combinations to find the most relevant data. NH will develop and refine these search terms with feedback from IM, CB, our information specialist. A draft strategy for Embase can be found in *Additional file 2*.

This search strategy was based on a sampling frame with the following focus:

*Context:* Older adults (60 years and above) from EMCs dealing with polypharmacy in primary care.

*Intervention:* Interventions aimed at optimising medicine use and the experiences of these older individuals, their family carers, and practitioners.

*Mechanisms:* identified from the programme theories.

*Outcomes:* quality of life, adherence, adverse events, disease symptoms, and patient satisfaction, as well as unexpected outcomes.

A formal literature search will include MEDLINE/PubMed, Embase, Scopus, Web of Science, the Cochrane Library, CINAHL, and PsycINFO. For grey literature, the ProQuest Dissertations & Theses Global, the King‘s Fund Library Database, NHS Evidence, and NICE will be searched. Additionally, we will use the snowballing technique to explore further papers, which could add contextual depth.

### Step 3 and 4: selection and appraisal of evidence

We will follow a systematic process of two steps to select and appraise articles using RAYYAN (a web-based application that helps facilitate the screening of titles and abstracts) (59). The first step will start by screening titles, and abstracts, against inclusion and exclusion criteria ***(Table 1)***. A 20% random sample will be checked by a second reviewer CB. Any disagreements will be resolved through discussion, or if necessary, by consulting with the rest of the team. For the second step, the documents which pass the initial screening will then undergo a full-text review to assess their relevance, rigor, and richness.

**Table 1:**
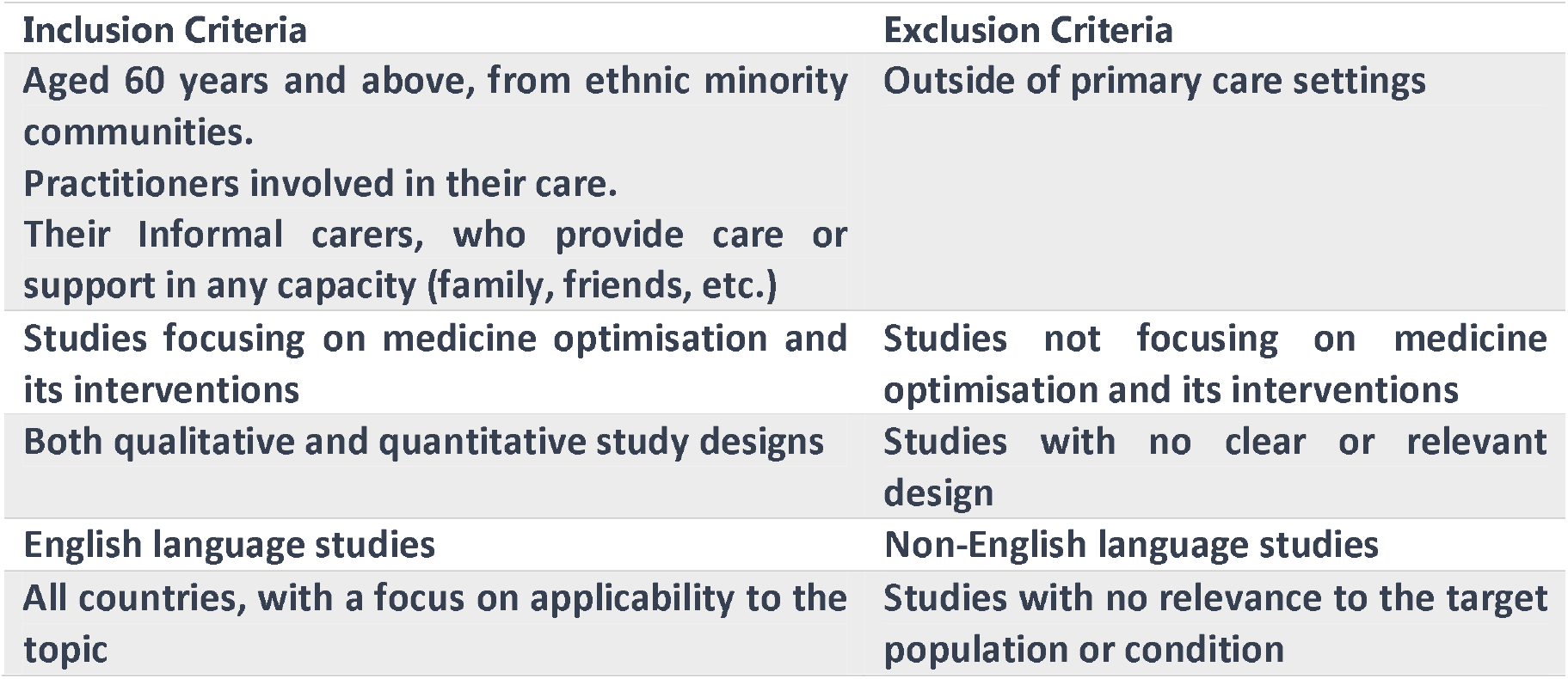
Inclusion and Exclusion Criteria.

The full-text articles will be critically assessed and rated on a scale of one to five stars in EndNote, reflecting their relevance, richness, and rigor in contributing to the development of the programme theory.

### Relevance

Relevance will be assessed by asking the following questions:

Do they provide insights or data into the challenges and considerations of medicine optimisation for older adults from EMCs dealing with polypharmacy in primary care?

Are they relevant to the development or refinement of program theories? (52)

We will use a relevance ranking system, which was used before in a previous study (60). With flexibility for studies outside the target age range (60 years and above) if they provide valuable insights:

- *High Relevance (4-5 Stars):* Direct insights into older adults from EMCs‘ medicine optimisation in primary care, specifically with polypharmacy. Studies including ages below 60 (e.g., 18-59) are considered if they offer significant, applicable insights.
- *Moderate Relevance (3 Stars):* Related insights into medicine optimisation, potentially applicable EMCs. Must provide considerable information that is relevant to the olders’ specific challenges.
- *Low Relevance (2 Stars):* General discussions on medicine optimisation or polypharmacy are not focused on the target older demographic or significantly younger populations.
- *No Relevance (1 Star):* Studies not aligning with the above criteria are excluded.

One-star ratings will indicate irrelevance, leading to exclusion, while two-star ratings will mark documents as uncertain, which will require a discussion with the second reviewer for potential re-evaluation. Three-star documents, although not directly contributing to the core theory, will provide useful background and therefore be included for context. Four-star and five-star documents will be identified as highly relevant and rich and will be integrated into the analysis, directly informing, and refining the programme theory.

### Rigor

Rigor is defined as the trustworthiness and credibility of the data (52). When evaluating studies based on rigor, we will look into the study‘s design, methodology, data collection, and analysis techniques (61). Documents that offer significant insights or potential contributions to theory development, even if not the most rigorously conducted, may still be included. However, a particular focus will be given to studies, that demonstrate a high degree of rigor.

### Richness

Richness refers to the depth and detail with which studies explore and present their findings and implications, especially concerning the lived experiences, challenges, and outcomes (62).

### Step 5: data extraction and synthesis

We will upload the full texts of the included papers into NVivo. This qualitative data analysis software tool will be used in coding relevant sections of the texts that relate to medication optimisation contexts, mechanisms, and their relationships to outcomes (63). The characteristics of each document (such as study objectives, methods, participant demographics, and key findings) will be systematically extracted and organised into an Excel spreadsheet.

The coding process within NVivo will include inductive, deductive, and retroductive approaches. Inductive coding will allow themes to naturally emerge from the data, which will ensure the analysis is grounded and responsive to the data itself (64). Deductive coding will involve applying pre-established codes based on the initial program theories, which will maintain alignment with the used theoretical framework (64). Retroductive coding will delve deeper, interpreting data of underlying causal mechanisms, thereby uncovering richer layers of understanding (65).

The data analysis will adopt a realist logic, which aims to make sense of the initial programme theories (52). This process will be supported by a series of critical questions to guide the analysis and synthesis of data, focusing on relevance, richness, and rigor. These questions will include interpreting the meaning of data in the context of the programme theory; making judgments about partial or complete CMOCs based on the data; and understanding how these CMOCs relate to the overarching programme theory. Also, interpretive cross-case comparisons will be employed to explain how and why certain outcomes occur, such as by examining differences in the levels of engagement in medication optimisation and cultural differences. Throughout this process, various forms of reasoning including juxtaposition, reconciliation, adjudication, and consolidation of data will be used (49). The SCT will be used again to help in explaining why certain pattern emerges from CMOCs.

## Discussion

The realist review aims to understand how medicine optimisation works/does not for older adults from EMCs with polypharmacy in primary care. This method will give a deep understanding of what works, for whom, and under which circumstances. Once we understand what is likely to work then we can start to develop effective interventions.

There are a number of limitations. Firstly, the inability to access the British Library‘s EThOS, due to technical issues which will restrict the inclusion of theses and dissertations which may offer unique insights. To mitigate this, we plan to use alternative platforms such as ProQuest Dissertations & Theses Global, which offer a collection of global dissertations and theses.

The review depends on English language studies, which could introduce a language bias (66) possibly overlooking significant research published in other languages (67). This limitation is important to recognise as relevant research may exist beyond English language publications.

Lastly, the findings of this review will provide insight into the context of specific primary care settings and the cultural context of the populations studied. Therefore, the generalisability of the results, beyond this context, may potentially be limited, and the conclusions drawn will need to be interpreted within the context of the reviewed studies (68). By following the RAMESES framework, we aim to improve the reflexivity and transparency of the process (52).

We will share the results of our review in line with the RAMESES reporting standards (52). This will include publishing the review in a scientific journal and submitting abstracts for presentation at both national and international primary care and pharmacy practice conferences.

## Data Availability

No data were produced in this study as it is a protocol. All relevant data and materials will be made available upon request once the study is completed.

## Declarations

## List of abbreviations

CMOCs Context: Mechanism, Outcome Configurations
EMCs: Ethnic Minority Communities
IPTs: The Initial Programme Theories
MO: Medicine Optimisation
RAMESES: Realist and Meta-narrative Evidence Syntheses: Evolving Standards

## Authors and Affiliations

Nesrein Hamed, College of Health and Life Sciences, University of Aston

Clare Bates, National Institute for Health and Care Research

Dr Muhammad Umair Khan, College of Health and Life Sciences, University of Aston

Professor Ian Maidment, College of Health and Life Sciences, University of Aston

## Ethics approval and consent to participate

Ethical approval is not required for this review, as it is a secondary data analysis that does not involve primary data collection from participants.

## Consent for publication

Not applicable.

## Availability of data and materials

Not applicable.

## Competing interests

None declared.

## Funding

This work is supported by HMMCT.

## Authors‘ contributions

NH worked on designing the review and drafting the manuscript.

IM, CB and MUK advised on the methods and edited and approved the manuscript.

## Authors‘ information (optional)

Twitter: @NesreinHamed @Prof_Ian_M @clarebates74

## Notes

### Competing Interest Statement

The authors have declared no competing interest.

