## Additional File 2 for "Understanding the Challenges of Medicine Optimisation Among Older People from Ethnic Minority Communities (Aged 60 Years and Above) With Polypharmacy in Primary Care: A Realist Review Protocol"

**Embase**

**Host:** Ovid

**Date range searched**: 1974 to present.

**Date searched:** 30/01/2024.

**Searcher**: NH

**Hits n:** 269

| | | |
|---|---|---|
| 1 | (Medic* Management or medic* optimi?ation or drug utili?aton review or medic* reconcil* or medic* review or structured medic* review or deprescri*).m_titl. | 12406 |
| 2 | (Medic* adherence or Medic* compliance or patient satisfaction or "Inappropriate prescribing" or Overprescrib* or "Medication burden" or "adverse events").m_titl. | 40928 |
| 3 | (older or elderly or aging or "old age" or "late life" or "60 years and above" or Geriatric*).m_kw, ab, titl. | 1976798 |
| 4 | (Middle East* or Afric* or Asia* or Caribbean or "West Indies" or Bangladesh* or China* or India* or Somali* or Ethiopia* or Nigeria* or Kenya* or Uganda or Syria* or Pakistan* or ethnic minorit* or Black or Asian or "people of colour" or Race* or "mixed race" or "mixed racial" or "Black British" or "indian subcontinent" or Gyps* or "irish traveller" or "African Americans" or "Asian Americans" or Blacks or "Hispanic Americans" or Arabi* or Hindu* or Hindi or Muslim or Islam* or Tamil* or Lanka* or Urdu or Bengali* or Emigran* or Immigran* or Refugee* or migrant* or asylum seeker* or BAME or BME). m_kw, ab, titl. | 2529523 |
| 5 | (Polypharmacy or multipl* medic* or multipl* drug* or many medic* or "many drugs"). m_kw, ab, titl. | 75407 |
| 6 | ("Primary care" or "Community health" or Family practi?e or General Practitioner or Pharmacy or GP or Family Medicine). m_kw, ab, titl. | 609399 |
| 7 | 1 or 2 or 5 | 126360 |
| 8 | 3 and 4 and 6 and 7 | 269 |
